# Collaboration and Innovation Patterns in Diabetes Ecosystems

**DOI:** 10.1101/2024.04.25.24306351

**Authors:** Odile-Florence Giger, Estelle Pfitzer, Wasu Mekniran, Hannes Gebhardt, Elgar Fleisch, Mia Jovanova, Tobias Kowatsch

**Author notes:** = shared last authorship (Mia Jovanova, Tobias Kowatsch).

## Abstract

**Background:** The global prevalence of diabetes is increasing and has stimulated new technological advancements in disease management. Although there are many digital health companies with a focus on diabetes, building them up at scale is difficult due to a heterogeneous, inefficient, and fragmented healthcare system. While ecosystems, or collaborative value creation, could help address system fragmentation; the current diabetes ecosystem remains not fully understood. Therefore, this paper analyzes the digital transformation of the diabetes ecosystem and deducts innovation patterns. We address the following research questions: (1) What are emerging organizations in the current diabetes ecosystem? (2) What are the value streams in the current diabetes ecosystem? (3) Which innovation patterns are present in the ecosystem?

**Methods:** We conduct a literature review and a market analysis to describe the organizations and value streams in the diabetes ecosystem, both before and after the digital transformation. We visualize the diabetes ecosystem using the e3-value methodology (RQ1 and RQ2). Next, expert interviews are conducted to validate the resulting diabetes ecosystem and deduce innovation patterns (RQ3).

**Results:** First, we show that the digital transformation gives rise to emerging organizations across eight segments: real-world evidence analytics, healthcare management platforms, clinical decision support, diagnostic and monitoring, digital therapeutics, wellness, online community, online pharmacy (RQ1). Secondly, we visualize the value streams between emerging organizations in the current diabetes ecosystem, highlighting the key role of patient data as currency (RQ2). Ultimately, we derive four innovation patterns in the current diabetes ecosystem (RQ3); namely open ecosystem strategy, outcome-based payments, platformization (connecting stakeholders), and user-centric software.

**Conclusions:** We demonstrate how traditional value chains in the diabetes ecosystem transition to platforms and outcome-based payment models, guiding strategic decisions for companies and healthcare providers. These innovation patterns may apply to similar ecosystems in other disease areas, aiding organizations in forecasting future dynamics.

## Introduction

Approximately 537 million people are suffering from diabetes today.^1^ Poor glucose control leads to many complications that reduce quality of life of patients, life expectancy and increase the healthcare cost of the disease.^2,3^ Total direct costs of diabetes worldwide are estimated at $966 billion in 2021, a 316% increase over the last 15 years.^1^ Hence, diabetes stands as a widespread, detrimental, and expensive condition.

Diabetes management is characterized by various approaches that often challenge the patients.^4^ Their tasks of self-management, which involve regular medication and insulin administration, frequent monitoring of blood sugar levels, strict dietary control, and regular exercise, can be challenging to manage.^5,6^ Many patients struggle with medication dosage, scheduling appointments, and their capacity to effectively handle numerous self-management responsibilities. ^7^ Successful management of diabetes depends heavily on patient adherence to specific behaviors. Poor adherence can result in morbidity, mortality, and poorer quality of life.^8^

At the same time, technological progress has brought many innovative approaches to enhance self-management for patients with diabetes. Among these, digital health technologies (DHTs), defined as “computing platforms, connectivity, software, and sensors [used] for health care and related uses.” ^9^ can improve access to health information for both patients and providers, enable remote patient monitoring, and deliver timely healthcare recommendations and reminders to patients.^10^ Therefore, DHTs for diabetes can support patients^5^ and providers.^11^

Nevertheless, new DHTs at the nexus of the healthcare and technology sector require navigation for a business model to be successful.^12^ But even if digital health business models are successfully built, scaling them up is often challenging due to heterogeneous, inefficient, and fragmented healthcare systems.^13^ It is, therefore, essential to consider the DHT transformation within the broader diabetes ecosystem. Studies to date often focus on *intra-organizational* perspectives, such as a single company’s products, processes, services, or business model, in isolation.^14,15^ However, research is lacking from a broader, *inter-organizational* level. This landscape view is critical as the digital transformation impacts the co-creation between multiple stakeholders in a larger ecosystem.^16^ Organizations need to understand the incentives and motivators of all ecosystem parties and align regulators, patients, physicians, providers, payers, and partners.^17^ Addressing this gap is a key step to develop solutions for a fragmented healthcare system. Therefore, this work aims to describe the current diabetes ecosystem, in light of the digital transformation, i.e., what are the emerging (or new) organizations, value streams and corresponding innovation patterns. To this end, the following research questions are formulated:

(1) What are the emerging organizations in the current diabetes ecosystem?
(2) What are the value streams in the current diabetes ecosystem?
(3) Which innovation patterns are present in the ecosystem?

### Theoretical Background

#### Ecosystems in healthcare and platform ecosystems

Studies^18^ identified three major research directions in the field of ecosystems, namely “Business Ecosystems,” “Innovation Ecosystems,” and “Platform Ecosystems.” Most current definitions of ecosystems ^18–23^ exhibit significant content overlaps, even though they are often formulated in different ways. According to prior work^24^ the essential components of these definitions encompass four elements, linking three operative concepts – interdependencies, networks, and self-interested actors – with the most common success criterion of an ecosystem: The collaborative value creation of actors in a manner that an individual actor would not be capable of achieving alone.

1. **Interdependencies:** To accurately depict an ecosystem, it is essential to recognize and analyze all alliances and relationships among the actors. A thorough examination of the intricate interactions within the ecosystem can lead to a more profound comprehension of the fundamental dynamics and dependencies at play.^19^
2. **Network:** The network of an ecosystem is defined by the structure of relationships among its members. Additionally, the network is characterized by members having defined positions and activity flows, and mutual agreement among actors regarding these positions and processes. ^19^
3. **Self-interested actors:** In essence, actors can be classified into one of three groups: orchestrators, members, and other organizations. Orchestrators^25^, also known as architects^18^ cornerstones ^22^, hubs ^23^, or sponsors ^20^ are organizations that possess the capability to both support and derive benefits from the success of an ecosystem through a combination of resources, leadership, and control.^20^ They play a crucial role in consolidating the industry by providing a dominant design for the ecosystem, comprehensively supporting the business model’s value proposition, facilitating collaboration among actors, and fostering collective innovation.^22^ The functions of *members* within an ecosystem have been inadequately explored in scientific research thus far. Although members of an ecosystem generally aim to promote the overall success of the ecosystem, their self-interest takes precedence.^26^ Tsujimoto et al.^23^ have also introduced other organizations in their depiction of the multi-actor network, which are classified as actors within an ecosystem. This includes regulators such as government organizations, regulatory authorities, and standard-setting bodies, associations, interest groups ^27^, nonprofit organizations (NPOs), universities, and individuals.^20^
4. **Value creation:** It can be stated that the raison d’être of an ecosystem always lies in the fact that actors collectively create value that they would not be able to achieve individually.^28^ An open network prioritizes overall value creation, offering compatibility with various products and established standards. In contrast, a closed network emphasizes individual actor value creation within the same product family, requiring market power and substantial investments.^22^

Specific to healthcare, prior work^29^ highlights the key role of platform ecosystems. A platform ecosystem is described as a ‘Hub-and-Spoke’ model, comprising a sponsor with a platform (’Hub’) and providers of complements (’Spoke’), which enhance the value of the platform for consumers.^18,23^ Innovation happens when platform owners want to expand their functionalities to external stakeholders with additional competencies., i.e., connecting disparate stakeholders.^30^ For example, prior work has shown that digital platforms orchestrate a platform-mediated ecosystem to co-create value with different partners and provide added value to patients, for example in the context of electronical medical records or telemedicine platforms.^30^ Together, platform ecosystems in healthcare are becoming increasingly common as digital technologies continue to transform the industry.

#### Ecosystem Analysis

To characterize an ecosystem, it is necessary to first identify existing relationships between actors, to better understand their underlying dynamics and dependencies.^19^ Different methods have been proposed to analyze and visualize ecosystems.^31^ Here, we chose the e3-value methodology^32^ which offers a structured framework for systematic identification, analysis, and visualization of multi-stakeholder relationships in healthcare contexts. The e3-valuemethodology is particularly well suited due to its’ conceptual modeling strength in capturing complex, multi-enterprise relationships and economic value exchanges among actors.^32–34^ The main aspects of the e3-value methodology can be described as follows:

1. **Actors:** An actor is recognized by its surroundings as an autonomous economic (and frequently legal) entity.
2. **Market segments:** A market segment divides a market into groups with common properties.
3. **Value objects:** Actors engage in the exchange of value objects, which can be services, goods, money, or even consumer experiences. The crucial aspect is that a value object holds significance for one or more entities.
4. **Value ports:** An actor utilizes a value port to indicate its intention to offer or seek value objects to its surroundings. The port concept allows abstracting from internal business processes and concentrates solely on how external entities and other components of the value model can be seamlessly integrated.
5. **Value interface:** Actors have value interfaces, which can be a single offering or involve both ingoing and outgoing offerings, reflecting economic reciprocity. This assumes actors offer something valuable if they receive appropriate compensation. The value interface models an actor’s willingness to give and receive, while a value offering indicates objects are requested or delivered in combination.
6. **Value exchange:** A value exchange is employed to link two value ports, signifying the mutual willingness of the connected actors to exchange value objects.

## Methods

To answer our research questions, we follow the e3-value methodology^32,35^, building on prior work.^30^ Our method consists of two steps. In step one, we conducted a literature review and a market analysis to identify existing organizations and value streams in the diabetes context, before and after the digital transformation. We distinguish between traditional diabetes ecosystem (before 2013) and current diabetes ecosystems (after 2013), due to a significant increase in articles discussing the digital transformation thereafter^36^ and given our specific aim to analyze the emerging organizations in response to the digital transformation. Thus, we visualize both ecosystems. In step two, we conducted expert interviews and applied these insights to refine our ecosystem visualizations and derive resulting innovation patterns (see Figure 1 for Study Workflow).

**Figure 1.**
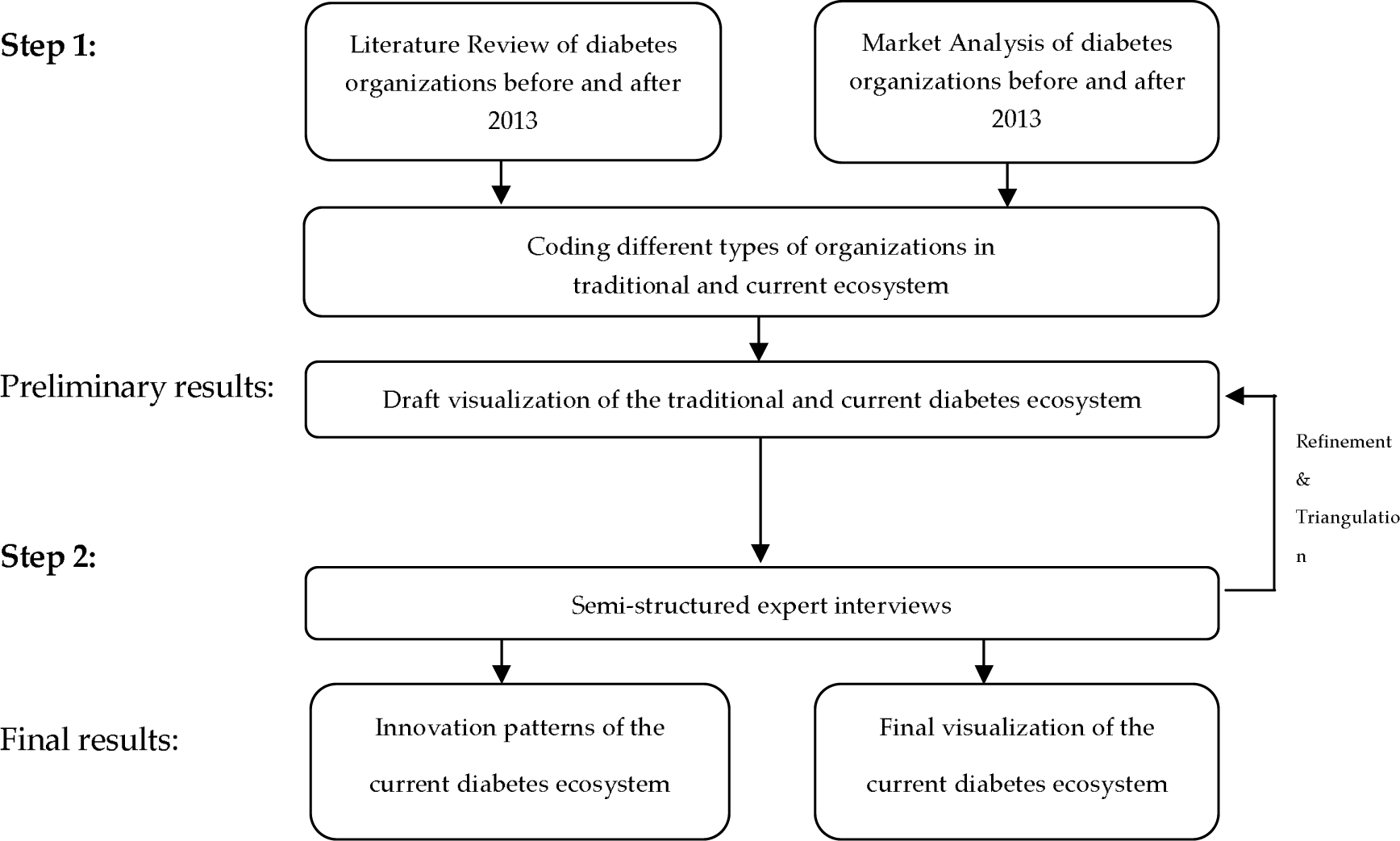
Study workflow and methodological process.

### Literature review

The literature review aimed to identify emerging organizations—those operating within the current ecosystem but not within the traditional one—and to map out emerging value streams. To this end, following prior work^37^, we conducted two separate literature searches, to capture the diabetes ecosystems both (a) before and (b) after the digital transformation, marked in 2013.^36^ The first search included empirical studies until December 31, 2012, while the second search started from 1 January 2013 – 30 September 2023. Based on prior work^30^, for the traditional diabetes ecosystem we used the following key terms: ((“Stakeholders” OR “Value network” OR “financial incentives”) AND (“diabet*”)). For the current diabetes ecosystem, we used the following: ((“stakeholders” OR “value network” OR “financial incentives”) AND (“diabet*”) AND (“digit*” OR “innovation”)). The final search included 14 articles for the traditional ecosystem and 31 articles for the current diabetes ecosystem. For more details on eligibility criteria, data sources, and search strategy, and paper selection see Appendix A.

### Market Analysis

The goal of the market analysis was to supplement the literature review with additional emerging organizations and value streams, not typically captured in academic literature.^30^ We conducted a search on diabetes-related companies in Pitchbook, a widely used platform in business science.^38,39^ Pitchbook enables the identification of organizations associated with the diabetes ecosystem as well as new technologies within the field. Consistent with our literature review, we conducted two separate market searches, to capture the diabetes ecosystems both (a) before and (b) after the digital transformation, marked in 2013. We applied the search terms “Diabetes” and “Diabet*” to both the traditional and current diabetes ecosystems. From these searches, we selected the top 100 organizations established before 2013 and the top 100 organizations founded after 2013, based on revenue^40^ with searches conducted on the 30^th^ of October 2023. For more details on company eligibility criteria see Appendix B.

### Literature review and market analyses thematic coding

We used a structured content analysis, including an inductive category development^41,42^ to code the emerging organizations and value streams, resulting from both the literature review and market analyses. To do this, we first coded the resulting organizations into pre-defined market segments and generic roles, following prior work.^30^ Specifically, for the emerging companies, we followed and adapted the categorization provided by the Digital Therapeutics Alliance, the main international body overseeing digital therapeutic interventions, as relevant to diabetes management.^9^ Following codebook development, two researchers coded organizations into market segments and generic roles, and we assessed coder consistency by calculated Cohen’s Kappa. We received a Cohen’s Kappa of 0.862, indicating high intercoder reliability.^43^ See Appendix C for codebook examples.

### Expert interviews and validation

We conducted additional expert interviews to provide feedback and validate assumptions about emerging organizations and value streams, derived from the literature review and market analyses.

Selection of interviewees and procedure

The expert interviews were conducted with healthcare experts of diabetes companies and healthcare providers using a semi-structured approach.^44^ These individuals were selected for having expertise in key market segments. See Appendix D for interviewee roles and domain expertise. Ten interviews were conducted online via video conference and one in person.

Interview coding and derivation of innovation patterns

Based on existing work^30^, we used a thematic analysis method^45^ to compare the traditional ecosystem with the current ecosystem and specifically derive innovation patterns. We define innovation patterns as new approaches in the current diabetes system that aim to solve reoccurring problems.^46^ Interview-derived innovation patterns help us complement secondary analyses by providing qualitative insights on complex ecosystem changes with potential transferability to other domains.^46–50^.

As such, we coded interview responses specific to innovation patterns, following these established steps in the field^45^: (1) familiarization with the data, (2) code generation, (3) theme generation, (4) theme review, (5) theme definition and labeling, and (6) identification of illustrative examples.

## Results

The results are structured along the three research questions: (1) emerging organizations, (2) value streams and (3) innovation patterns in the current diabetes ecosystem.

### Emerging organizations and value streams in the current diabetes ecosystem

The literature review and the market analysis revealed several emerging organizations from the traditional to the current diabetes ecosystem. Here we discuss these organizations (RQ1) with respect to their generic roles and value streams (RQ2). See table 1 for the generic roles and market segments of the emerging organizations in the current ecosystem, and see Appendix E for organizations in the traditional ecosystem.

**Table 1.** Emerging organizations in the current diabetes ecosystem.

| Generic role | Market Segment | Description |
| --- | --- | --- |
| Digital Health Technology - Industry & Admin facing | General definition | Digital health solutions for non-hospital/ health system stakeholders (e.g., pharma, MedTech, payors etc.): Clinical administration and management tools, predictive analytics, clinical trial management.<sup>9</sup> |
|  | Real world evidence analytics | Real world data (RWD) and real-world evidence (RWE) plays an increasingly important role in clinical research since scientific knowledge is obtained during routine clinical large-scale practice and not experimentally as occurs in the highly controlled traditional clinical trials. Real world evidence data can be used for biomarker discovery or validation, gaining a new understanding of a disease or disease associations, discovering new markers for patient stratification and targeted therapies, new markers for identifying persons with a disease, and pharmacovigilance.<sup>52</sup><br>**Example:** Glooko, Hedia |
| Digital Health Technology - Healthcare provider facing | General definition | Platforms and Health Information Technology and digital health solutions that supports clinicians with managing their patient population.<sup>9</sup> |
|  | Healthcare management platform | A healthcare management platform is an online platform designed to enhance collaborative efforts among health care providers, facilitating efficient sharing of data.<sup>53</sup> Furthermore, it can encompass telehealth, which gathers, transmits, and communicates patient's personal health information to their healthcare provider or extended care team, all while being conducted beyond the confines of a hospital or clinical setting, typically in the patient's home.<sup>54</sup><br>**Example:** Sirma |
|  | Clinical decision | A clinical decision support system aims to enhance healthcare delivery by integrating targeted clinical knowledge, patient |
|  | support | data, and additional health information to improve medical decision-making.<sup>55</sup><br>However, there is a growing trend of developing systems with the ability to utilize data and observations that would otherwise be inaccessible or incomprehensible to humans. Artificial intelligence methods in combination with the latest technologies, including medical devices, mobile computing, and sensor technologies, have the potential to enable the creation and delivery of better management services to deal with diabetes.<sup>56</sup><br>**Example:** Hedia |
| Digital Health Technology - Patient-facing | Diagnostics & Monitoring | Diagnostics: “Validated digital tools for detecting and characterizing disease, measuring disease status, response progression, or recurrence. They are intended to support patient self-management of a specific diagnosed medical condition through education, recommendations, and reminders.”<sup>9</sup><br>Monitoring: “Solutions intended to monitor specific patient health data that may be used to inform management of a specific disease, condition, or health outcome.”<sup>9</sup><br>E.g. Digital diagnostics, Digital biomarkers, Remote patient monitoring tools, Wearables and biometric sensors, Medication ingestible sensors<br>**Examples:** BeatO, TwinHealth |
|  | Digital Therapeutics | “Health software intended to treat or alleviate a disease by generating and delivering a medical intervention that has a demonstrable positive therapeutic impact.”<sup>9</sup><br>**Example:** VirtaHealth, Fitterfly |
|  | Wellness | “Disease-agnostic solutions that capture, store, and sometimes transmit health data and promote general well-being and healthy living “.<sup>9</sup> E.g. Lifestyle and wellness apps (Meal tracker, weight tracker, bolus calculator), Activity and fitness trackers, Wearables<br>**Examples:** CashWalk, ModifyHealth |
| Pharmacy | Online pharmacy | Online pharmacies are businesses operating on the internet that offer pharmaceutical products, including prescription medications, through online ordering and delivery services.<sup>57</sup> |
|  |  | **Examples:** Zur Rose, Doc Morris |
| Community | Online community | In a Diabetes Online Community members help each other out, educate each other, and share the steps they take every day to stay healthy while living with this very serious condition.<sup>58</sup><br>**Examples:** Diabetes Sisters, TuDiabetes |

The findings suggest that in the digital transformation of the diabetes ecosystem three generic roles emerged among organizations (*DHT – Industry & Admin facing, DHT -Healthcare provider facing, DHT – patient-facing*), covering eight market segments (real-world evidence data analytics, clinical decision support, healthcare management platform, wellness, diagnostic & monitoring, digital therapeutics, online community, online pharmacy). While there isn’t always a one-to-one mapping between each organization, role and market segment (i.e., companies may simultaneously cover multiple market segments), we follow this framework to synthesize and understand the diverse range of organizations. We explain these emerging organization in further detail next.

First, *DHT–Industry & Admin facing* companies emerged as important players. These refer to digital health solutions for non-hospital/ health system stakeholders (e.g., pharma, MedTech, payors etc.), namely clinical administration and management tools, predictive analytics, clinical trial management.^9^ One prominent example is Hedia (Interviewee 5). Hedia functions as a personalized diabetes assistant, leveraging artificial intelligence. It recognizes patterns and behaviors unique to each individual with diabetes, utilizing this information to provide tailored insulin recommendations, thereby enhancing the effectiveness of insulin treatment for the person managing diabetes.^51^ *DHT–Industry & Admin facing* companies like Hedia highlights the role of patient personalization and integration with patient management platforms, such as Glooko. With respect to value streams, these companies generate revenue by selling and interpreting personalized data, thus highlighting the importance of patient data as currency. For example, the data collected through Glooko is analyzed by Hedia and sold back to pharmaceutical or medical device companies.

Second, *DHT -Healthcare provider facing* primarily address healthcare providers and manufacturers such as medical device companies and pharmaceutical companies. These patient management platforms, such as Glooko, play a crucial role in connecting the entire diabetes ecosystem by bridging different stakeholders (medical device and pharmaceutical companies, health care professionals, and digital health companies). In another example, software companies like Sirma support healthcare providers in building up their digital practice through telemedicine or remote patient monitoring. These companies allow different medical specialists such as clinicians, nutritionists, psychologists, and/or endocrinologists to access the patient’s information in real-time, particularly as diabetes management requires a comprehensive support from a team of experts (Interviewee 9). By enabling real-time access to patient information across different specialties, these platforms streamline diabetes management and promote coordinated care among the patient’s entire medical team. Here, value is exchanged by providing software for healthcare providers and promoting compatibility between disparate stakeholders (Interviewee 5).

Third, when it comes to companies in the field of *DHT – patient-facing*, numerous companies aim to provide support and guidance directly to patients. These emerging organizations, such as digital therapeutics and patient monitoring companies like the virtual clinic Virta Health, focus on promoting lifestyle changes to manage type 2 diabetes. Additionally, other companies operate within the wellness domain, like CashWalk, which aims to motivate individuals to increase their physical activity levels. Importantly, these applications are not exclusively for diagnosed patients; they can also be utilized by individuals without diagnosis from a preventative standpoint. In these contexts, companies often adopt a direct-to-consumer approach, where customers may pay out of pocket for services. Companies may create revenue by direct payments made by patients, patient data sharing, or by reimbursement from insurance providers.

### Visualizing organizations and value streams in the current diabetes ecosystem

Figure 2 shows the current diabetes ecosystem, namely connecting DHT companies with additional stakeholders (i.e., patients, healthcare providers, health insurances, regulators and government, medical device and supply companies, pharmaceutical and biotech companies, pharmacies, social support groups, laboratories, research centers, and wholesale industries). The general roles of the stakeholders are presented in gray (e.g. (*DHT – Industry & Admin facing, DHT -Healthcare provider facing, DHT – patient-facing)*, while the specific market segments are presented in white. Further, the emerging organizations are presented in dotted, white rectangles.

**Figure 2.**
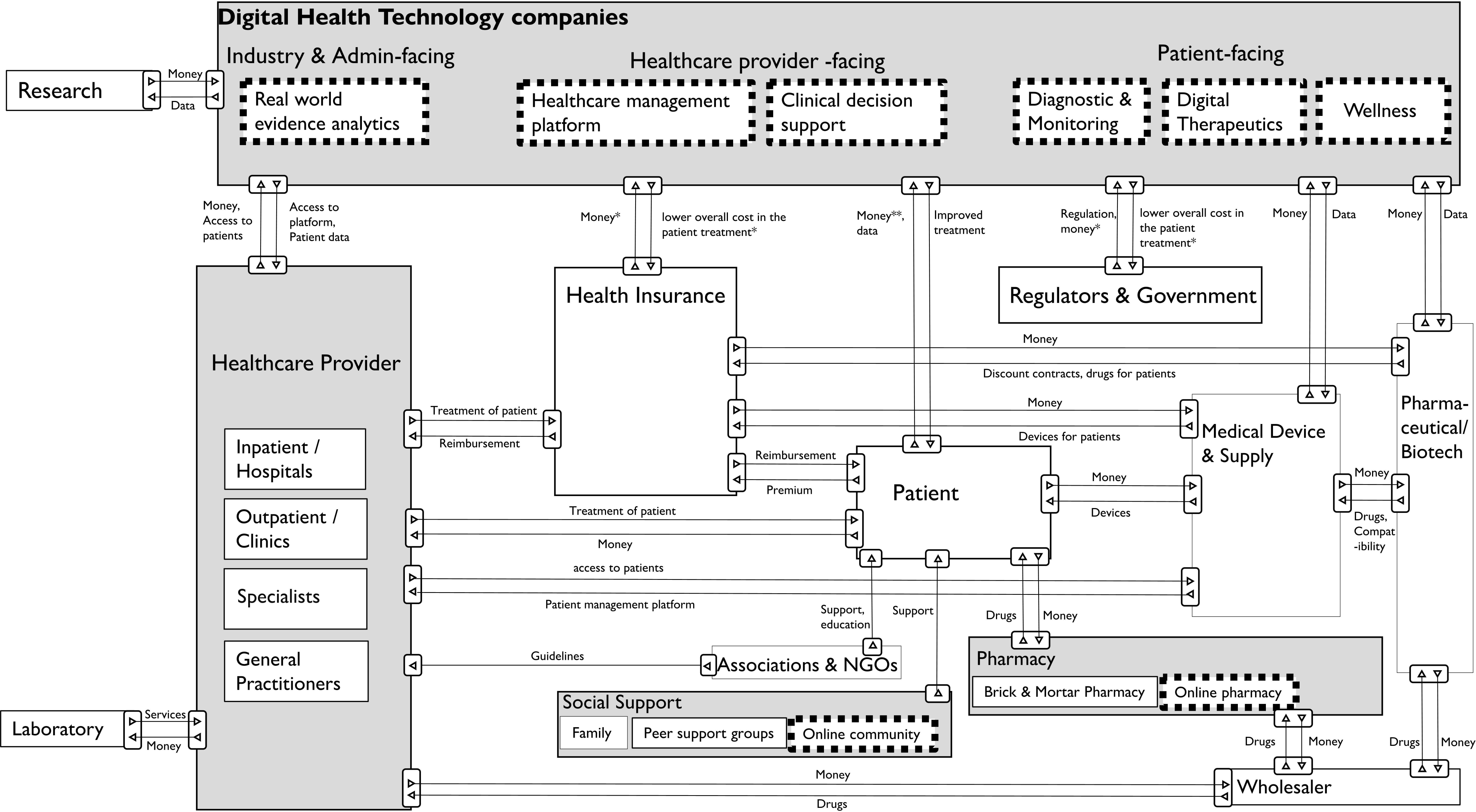
Visualization of the current diabetes ecosystem, derived from literature review, market analyses and expert interviews.

### Four innovation patterns in the current diabetes ecosystem

Expert interviews revealed four key innovation patterns in the current diabetes ecosystem, following the digital transition from the traditional diabetes ecosystem.

#### Innovation pattern 1 – Open Ecosystem Strategy: Pharmaceutical and medical device companies must consider participating in an open ecosystem

Pharmaceutical and medical device companies apply an open or closed ecosystem approach. In an open ecosystem, the goal is to encourage collaboration with all market participants, ensuring the compatibility of numerous devices with the platform. This enables patients to utilize devices from various manufacturers, granted these manufacturers have established partnerships with the digital platform (Interviewees 3, 4, 5, 7). Companies following this approach try to gain market access through different platform collaborations. This approach has two main objectives: firstly, to establish an open ecosystem by offering a variety of devices on the platform, thus promoting widespread adoption among healthcare providers, and secondly, to boost the sales of their products within the open ecosystem by influencing preferences toward those products (Interviewee 7).

Additionally, these device manufacturers participate in the open ecosystems of independent patient management platforms (Interviewees 2, and 5). The open ecosystem is generally more scalable and contributes to the growth of participating companies (Interviewees 5 and 7). Smaller companies need to join open ecosystems because they lack the influence to independently get healthcare providers to adopt their platforms. (Interviewee 7). However, a disadvantage is the integration costs that the device manufacturer must bear when connecting to an independent platform to ensure compatibility. Additionally, device manufacturers lose control over the data, as it can only be repurchased for a monetary amount (Interviewee 7).

In contrast, medical device and pharmaceutical companies in closed ecosystems create platforms that are only compatible to some degree with other companies (Interviewees 5 and 7). An example mentioned is Medtronic, which does not have as many collaborations as other companies, such as Dexcom. Companies choosing a closed ecosystem approach often have a big market share and are popular with healthcare providers (Interviewees 3, 4, and 5). Healthcare providers determine the worth of the ecosystem (Interviewee 6) and have the authority to decide which platform to choose (Interviewees 3, 4, and 5). If no healthcare provider adopts the platform, the ecosystem lacks value (Interviewee 5).

When a healthcare provider chooses to work with a specific manufacturer’s closed platform, they typically limit themselves to prescribing devices exclusively from that manufacturer. The reason is that devices from other manufacturers often don’t work with this closed platform. So, by choosing the manufacturer-dependent platform, the healthcare provider inadvertently excludes devices from other brands, favoring the manufacturer’s products (Interviewees 3, and 7). This means the manufacturer actively influences healthcare providers to prefer and prescribe their devices to secure a stronger market position for their products. The advantages of a closed ecosystem are less cybersecurity issues (Interviewee 9). A drawback arises from being generally less technologically advanced. This is because independent software companies like Glooko can focus all their resources on developing and improving their platform (Interviewees 3, 5, and 7). Furthermore, manufacturers of the closed platform must be capable of offering all classes of devices, even though historically they have only offered one device. This results in a lack of competitiveness for their devices at the device class level (Interviewee 5). Additionally, providing their platform involves a high resource investment (Interviewees 3, and 5). Both approaches showed advantages and disadvantages. Nevertheless, interviewees largely supported an open ecosystem strategy due to potential benefits for diverse stakeholders.

#### Innovation pattern 2 – Outcome-based payment: Organizations should be aware of new outcome-based payment as a revenue model and push it further

The evolving healthcare landscape anticipates a discernible shift toward outcome-based payment models as more patient data is collected. In this paradigm, pharmaceutical companies, medical device manufacturers, and healthcare providers will be required to substantiate the efficacy of their treatments. Using real-world evidence data facilitated by an open ecosystem is pivotal in enhancing treatment approaches. This data not only aids in continually refining treatment methodologies but also positions stakeholders with a stronger negotiating stance. Even in the present scenario, demonstrating the effectiveness of treatment affords these entities greater bargaining power, enabling them to secure more favorable pricing agreements with regulatory bodies. This forward-looking approach not only underscores the significance of real-world evidence but also emphasizes the strategic advantage in substantiating the value of patients’ health data and the importance of well-connected platforms (Interviewees 2, 4, 8, and 9).

Nevertheless, it was mentioned that in the case of diabetes, it’s quite challenging to adopt outcome-based treatment. This is because diabetes is a complex disease, making it difficult to pinpoint the specific effects of different interventions on patient outcomes (Interviewee 9). In the diabetes ecosystem, coordination incurs costs that prompt a dual assessment—financially and in terms of patient impact. The critical inquiry revolves around the cost-effectiveness of coordination efforts, weighing monetary investments against potential advancements. Simultaneously, it questions whether such coordination enhances patient experiences and outcomes in diabetes management. Striking a balance between these considerations is crucial to ensure that incurred costs translate into meaningful returns, both financially and in the overall well-being of patients (Interviewee 7). Another potential risk of new DHTs and medical devices is that they will be described to patients who do not need them. Healthcare providers might adopt the mentality of *«If you have a new hammer, every problem looks like a nail.”* In this case the diabetes ecosystem would get very expensive. Therefore, it was advised to only use new treatment possibilities if it also substantially benefits the patient (Interviewee 10).

#### Innovation pattern 3 - Platformization: Organizations should be aware of the platformization trend, decide on their role in the ecosystem and prioritize patient data as a key resource

In the context of open ecosystems, platform companies play a crucial role. An example is Glooko, which operates as an orchestrator, connecting different stakeholders such as healthcare providers, manufacturers, and patients. In some countries, the state or health insurance even pushes these platform ecosystems. For example, Sweden and Norway partly fund Glooko, whereas, in France, Glooko is paid by health insurance (Interviewee 5).

These platform business models work because more and more stakeholders in the ecosystem are interested in patient data. Patient data fuels innovation in healthcare as outlined by Interviewee 9: *“Patient data is a key resource in the diabetes ecosystem and can be treated as the new oil.*” Analyzing large datasets can lead to insights that drive medical research, treatment development, and the improvement of healthcare services. Companies and healthcare organizations recognize the monetary potential of leveraging this data for various purposes, including research, drug development, and targeted marketing. With the help of large data sets, actors can demonstrate the effectiveness of their intervention in improving patient health, also known as outcome-based treatment (Interviewees 2, 8, and 9). While manufacturers strive more and more in this direction, not all healthcare providers might favor it as they could fear transparent data comparing the effectiveness of treatments of different clinics (Interviewee 9). Cybersecurity plays a critical role in safeguarding patient data, ensuring the integrity of medical records, and protecting against the rising threat of cyberattacks. The confidentiality and privacy of patient information are paramount, and robust cybersecurity measures are essential to maintain trust in healthcare systems. (Interviewee 8, and 9). Striking a balance between robust cybersecurity practices and the openness of an ecosystem is crucial to ensure the advancement of healthcare technologies while maintaining the highest standards of data security and patient privacy (Interviewee 9).

#### Innovation pattern 4 - User-centric software: Organizations creating software must prioritize healthcare providers and patients with integration possibilities and user-friendly interfaces

Interviewees concluded that general practitioners, specialists, and other healthcare providers play a crucial role in the diabetes ecosystem, particularly as they often decide what medical device and drug is most suitable for a given patient (Interviewees 4, 5, and 8).

In the past, healthcare providers were often swayed to use medications from specific companies through incentives like kickbacks. However, in today’s competition-heavy landscape, companies need new strategies to encourage general practitioners to adopt their devices. One common tactic involves user-centric software solutions for patient management. Interestingly, there is still ample opportunity for new entrants to the market, especially those offering software with interfaces tailored to the preferences of general practitioners (Interviewees 4, 5, and 8).

Moreover, software must integrate seamlessly with various devices, simplifying the daily workflow for practitioners. Currently, healthcare providers grapple with many disparate medical devices, used by different patients. Navigating many different login procedures and interfaces is often described as inefficient, time consuming, and burdensome. This is why platform solutions like Glooko create value for healthcare providers (Interviewee 10). Thus, to attract healthcare providers, companies must build user-centric, integrative platforms that are accessible and intuitive (Interviewees 4, 5, 8, and 10).

Diabetes companies are also experiencing a shift towards consumerization (Interviewee 8). Patients increasingly gather personal data on their wearable devices, such as smartwatches and Fitbits. As data becomes more available in daily life, patients have more transparency on their disease management and health status (Interviewee 10). With this, there is a need for more personalized, convenient, and accessible data, prompting companies to adopt an open (vs. closed) ecosystem approach and encourage collaboration in response to this evolving landscape (Interviewee 8).

## Discussion

In this study, we conducted a literature review and a market analysis to describe the organizations and value streams in the diabetes ecosystem, both before and after the digital transformation. We first demonstrate the emergence of organizations across eight segments within the diabetes ecosystem: real-world evidence analytics, healthcare management platforms, clinical decision support, diagnostic and monitoring, digital therapeutics, wellness, online community, and online pharmacy (RQ1). Following this, we visualize the flow of value streams between these emerging organizations, highlighting the significance of patient data as a medium of exchange (RQ2). Finally, we uncover four distinct innovation patterns within the current diabetes ecosystem (RQ3): open ecosystem strategy, outcome-based payments, platformization (interconnecting stakeholders), and user-centric software.

When it comes to different market segments, especially monitoring technologies, and digital therapeutics emerged.^59^ Diabetes monitoring devices generating data around blood glucose pose an important opportunity. Monitoring devices promise to detect real-time health conditions by enhancing connectivity and integration, offering enhanced health efficiencies.^60,61^ Besides diabetes monitoring, it has been found that peer connections where patients can share their expertise in online communities can be a great opportunity to improve their condition.^62^

Regarding innovation patterns, results show that the diabetes ecosystem is moving more toward platformization.^63^ This can be confirmed by prior work which states that the healthcare industry is moving from linear value chains to two-sided markets and, now, to a multi-sided market mediated by platforms. As mentioned earlier, a platform ecosystem is described as a ‘Hub-and-Spoke’ model comprising a sponsor with a platform (‘Hub’) and providers of complements (‘Spoke’). This means that many organizations are transitioning from a traditional business model to a model where they act as a platform or intermediary connecting multiple parties who exchange value with each other.^18,64^ Today, we’re witnessing a rise in interconnected networks of digital technologies, information systems, and processing tools. These networks require a high level of interdependence among competencies and technological complementarity.^65–67^ Nevertheless, this process may be slow due to regulation and the general complexity of the healthcare system. Also, from the patient’s perspective, there are not yet many diabetics who use platforms like Glooko. However, this may change (Interviewee 9 and 10). Especially, patients can benefit from such a platform as it could make advice between patients and specialists easier and more efficient.^68^

Next, we found that manufacturers chose an open or a closed ecosystem approach. Both approaches showed advantages and disadvantages. Nevertheless, more interviewees believed in an open ecosystem strategy due to its potential benefits for many stakeholders. Also, a study analyzing ecosystems concluded that the strength of a company does not solely rely on its capabilities but rather on its ability to connect to various competencies. The capability to establish connections with competencies, the fundamental skill of network orchestration, is just as crucial as firms’ specific capabilities.^69^ From the standpoint of a platform company, making decisions about openness and control can be complex, especially in digital health, where several national, regional, and local governmental bodies are involved. Platform companies must strike a delicate balance between leveraging boundary resources to open the platform to external actors and maintaining control over third-party innovations in the periphery. These boundary resources include but are not limited to application programming interfaces, software development kits, contract agreements, app distribution channels, and similar tools that enhance the value for third-party developers.^29^ Sometimes, governments force companies to share their platforms. For instance, the EU is passing new laws, such as the Digital Markets Act, to ensure fair competition online. Apple’s App Store is being watched closely for how it controls app distribution. To follow these laws, Apple is changing its App Store rules in Europe, giving users and developers more freedom and access. This change is meant to encourage competition and new ideas in online markets.^70^ To navigate the complex decision-making structure of an ecosystem, studies^71^ highlight using a National Architecture Framework as a coordination mechanism. This framework provides application programming interfaces and guidelines to facilitate the development of third-party modules across the platform ecosystem. This underscores the multitude of actors in the health domain, spanning both the platform core and periphery.^71^ Especially, the government faces the task of bridging the gap between promoting an open ecosystem for the societal benefits of extensive health data exchanges and ensuring the security and protection of individual health data.^72^

We also showed that patient data is increasingly important, as currency, in the diabetes ecosystem. Other studies found that integrating more patient-generated healthcare data could lead to novel opportunities for innovation^73^ for example, providing personalized treatments and predicting treatment outcomes.^74^ Applying artificial intelligence to this data can further push innovation.^75^ Although patient data encompasses a lot of promising opportunities, it also has drawbacks. Sharing patient data with different organizations is complex due to privacy, cybersecurity, and legal issues.^69^ Organizations must ensure that patient data is used safely and ethically as technology advances.^69^ Furthermore, to further promote digital health technologies an ex-ante and ex-post evaluation of platform organizations should be conducted.^68^

From a healthcare provider perspective, organizations must tackle the concerns related to the hesitancy of healthcare providers to share their data transparently. This is essential for successfully implementing outcome-based reimbursement models (Interviewee 9). Furthermore, we found that healthcare providers prefer systems that are easy to use. This is also supported by other researchers^76^, who found that healthcare providers prefer tools that are simple to set up and interoperable.

We contribute to the ecosystem theory in three ways. Firstly, by analyzing the diabetes ecosystem, we could present empirical evidence of how traditional value chains transform into platforms, connecting different stakeholders. This transformation has also been described by others.^18^ Our findings confirm existing theory^28^ that actors in an ecosystem want to create value collectively that they would not achieve individually. This is shown by the example of medical device and pharmaceutical companies wanting to cooperate with DHT companies. Secondly, we enhance the literature of platform ecosystems in healthcare^30^ with empirical evidence of a platformization trend in the field of diabetes. We can add to the existing theory that there is a tendency towards an open ecosystem approach that has been pushed by the platformization trend. Lastly, we confirm the theory that digital platforms create value by providing technological components utilized by complementors to create new products or services.^77,78^ This was shown by Glooko, that leverages their algorithm to interpret and resell the patient data to pharmaceutical and medical device companies.

For practitioners, leveraging the visualization of the diabetes ecosystem can be instrumental in facilitating strategic collaboration options and conducting competitive analyses. This aids in crafting a tailored strategy aligned with the company’s objectives. Moreover, it underscores the significance of the platformization trend, prompting thoughtful consideration of how to engage with it effectively. For emerging companies and startups, this ecosystem visualization helps illustrate the diverse incentives of various actors and how they can potentially generate revenue through their digital services. Additionally, the paper enlightens healthcare providers on their decision to participate in an open or closed ecosystem, emphasizing the impact on the patient. Policymakers and health insurers gain insights into ecosystem dynamics.

### Limitations and outlook

This study has several limitations. First, we have created different generic roles and market segments based on a literature review and a market analysis. Although several authors were involved in that process and validated these categories with interviewees of the diabetes ecosystem, others could end up with a different categorization of these organizations. Second, we assessed 200 companies in the market analysis. Analyzing more companies could bring up a more granular view of the diabetes ecosystems and their market segments. Third, most of the interview participants are from Europe. While most interviewees held international positions, experts from other continents and countries might see the diabetes ecosystem differently. Many Asian organizations could not be assessed in detail due to the language used on their website. Fourth, we generalized the visualization of the diabetes ecosystem to a high degree. The value streams might be slightly different in other countries. However, this visualization aimed to understand the dynamics of globally operating organizations from a broader perspective. This is why we ended up with a rather generic visualization.

Exploring the dynamics of open and closed ecosystems could be a promising avenue for future research, unraveling the optimal strategy for different actors in varied contexts. Researchers should investigate crafting an ideal blueprint for open ecosystems, identifying essential components for their success. Additionally, understanding the pivotal role of various actors, such as the state or health insurance, in driving this paradigm is crucial for comprehensive insights. Furthermore, it becomes imperative to ascertain the potential cost savings associated with adopting an open ecosystem approach and thoroughly examine its tangible benefits for the patient’s well-being. Lastly, it should be assessed if, in other chronic disease areas, similar ecosystem structures and innovation patterns evolved or will develop soon and to what extent the patterns of the diabetes ecosystem are unique.

## Conclusions

Overall, we examined the global diabetes ecosystem, illustrating the shift from traditional value chains to data-driven platforms and outcome-based payment models in response to digital transformation. Our findings contribute to ecosystem theory and provide strategic insights for diabetes organizations to plan proactively.

## Data Availability

All data produced in the present work are contained in the manuscript

## Appendix A

### – Literature Review

**Table 2.** Inclusion criteria and exclusion criteria.

| Inclusion Criteria |  | Reason for inclusion |
| --- | --- | --- |
|  | Research focus | Studies that identify (two or more) stakeholders and value streams in the field of diabetes |
|  | Year | First Literature review: Studies until 31/12/2012<br>Second literature review: studies from 01/01/2013 until 30/09/2023 |
|  | Language | Only English studies are considered |
|  | Publication type | Peer reviewed (qualitative and quantitative) literature |
| Exclusion Criteria |  |  |
|  | Research focus | Studies that do not identify or discuss (two or more) stakeholders in the field of diabetes. |
|  | Publication type | Grey literature (news articles, company publications, annual reports, NGO studies, presentations, catalogues) |

#### Data Sources

We used the same databases as related work, namely EBSCOHOST, Emerald Insight, IEEE Xplore ACM Digital Library.^30^ These databases are widely acknowledged and employed for conducting studies in the field of business and healthcare.^79,80^ We further adapted the databases selection to avoid imbalances in the number of study results between the traditional and current diabetes ecosystems. Thus, for the traditional diabetes ecosystem, we used EBSCOHOST and Emerald Insight. For the current diabetes ecosystem after 2013, we additionally included IEEE Xplore, ACM Digital Library and PubMed, as these databases help capture intersection of computer science, life sciences, and business, particularly in information systems and technology-driven business models to help capture emerging organizations.^30^

#### Search Strategy

We scanned the abstracts and titles of the studies across databases. See table 3 for key terms for the traditional diabetes ecosystem and table 4 for key terms for the current diabetes ecosystem:

**Table 3.** “Traditional” Diabetes ecosystem before 2013.

| Keywords used | EBSCOHOST | Emerald Insight | Total |
| --- | --- | --- | --- |
| “stakeholders” AND “diabet*” | 79 | 6 | 85 |
| “value network” AND “diabet*” | 74 | 95 | 169 |
| “financial incentives” AND “diabet*” | 48 | 23 | 71 |
| Total articles | 201 | 124 | 325 |

**Table 4.**
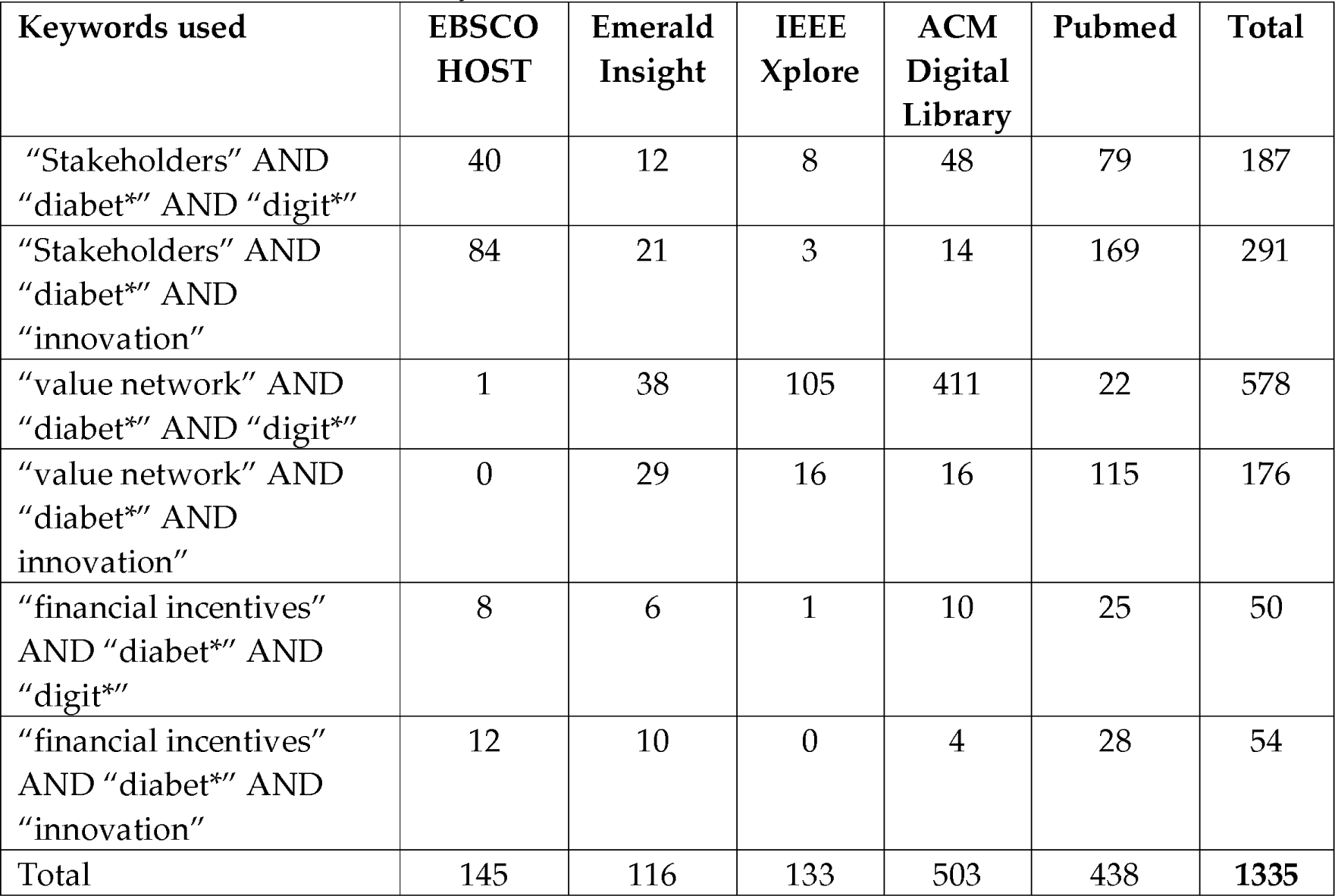
Current Diabetes ecosystem after 2013.

#### Paper Selection

Our study selection process follows the Preferred Reporting Items for Systematic Reviews and Meta-Analyses (PRISMA) flowchart and is depicted in figure 3.^37^ Following standards in the field^81^, two researchers (OFG and EP) conducted the paper selection process described as follows. The authors individually reviewed the abstracts of all papers and categorized them into three categories (A: relevant for the research objective, B: relevance unclear initially, C: not relevant). The authors compared and discussed their categorizations, addressing any unclear discrepancies, and only papers categorized as A were selected for analysis. Finally, 14 papers were selected as eligible for the “traditional” diabetes ecosystem up to 2013 and 31 papers were deemed eligible for the current ecosystem after 2013.

**Figure 3.**
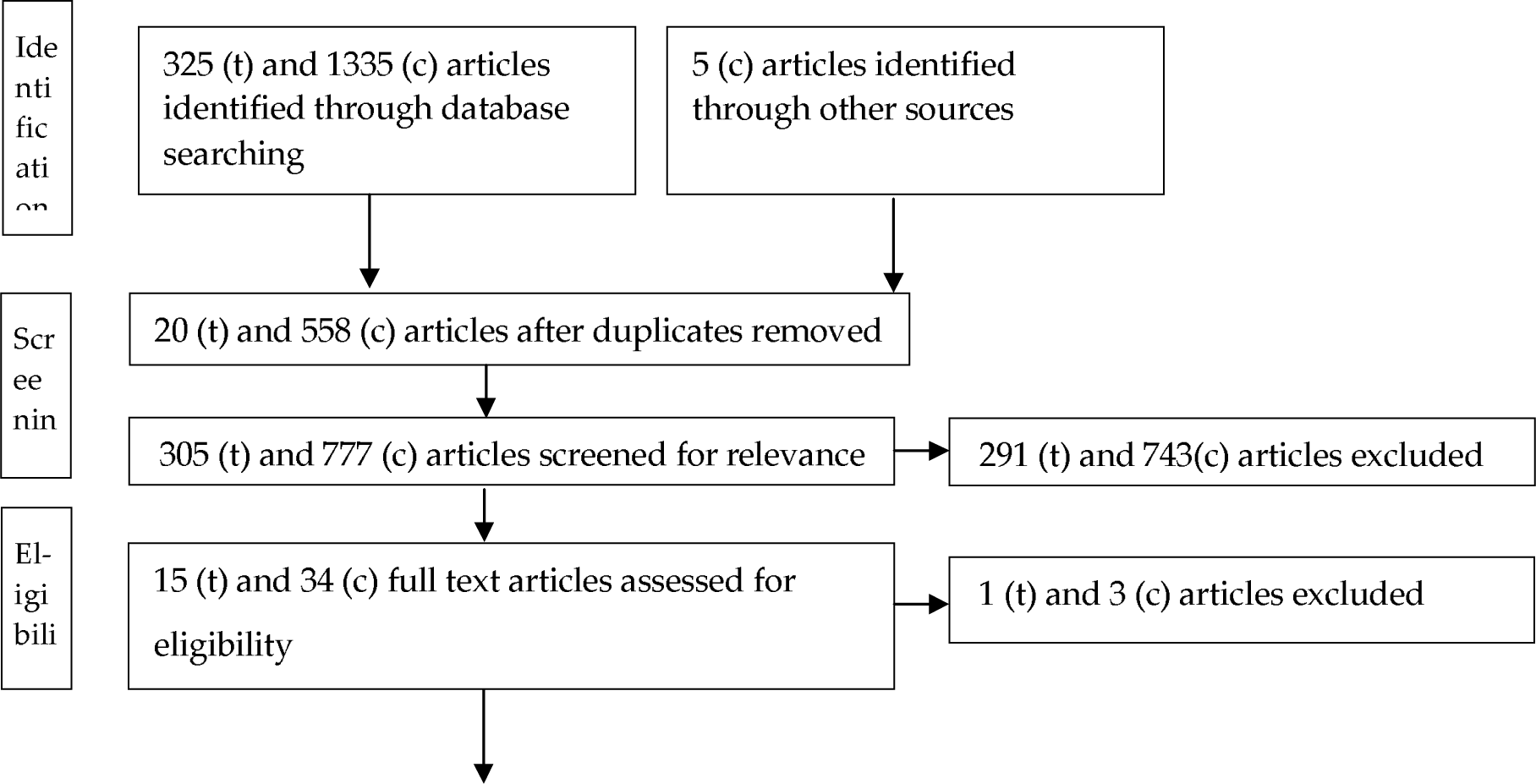
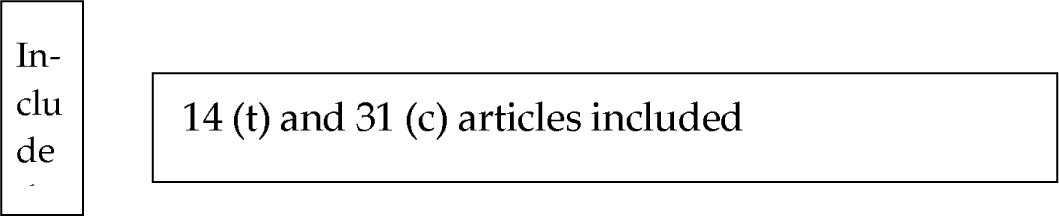
Flowchart of the literature review. (t) stands for articles from the “traditional” ecosystem, whereas (c) stands for the current ecosystem.

#### Procedure for thematic analysis and ontological organization

We conducted an inductive thematic analysis. This approach requires reading and re-reading data in iterative cycles to identify themes and categories. In this process, we developed superordinate classifications containing subclasses of diabetes organizations and checked for duplication and redundancy at each level.^82^ We prepared a table where we compared the coding of the authors. The table contained the following information: title, abstract, authors, publication date, actors, market segment, and description of these actors. Two researchers (OFG and EP) scrutinized each articles individually to identify the market segments, actors and value streams. The third author (WM) re-assessed and resolved the discrepancies.

## Appendix B

### – Market Analysis

#### Eligibility criteria

The inclusion and exclusion criteria are outlined in table 5.

**Table 5.** Inclusion and exclusion criteria.

| Inclusion criteria |  | Reason for inclusion |
| --- | --- | --- |
|  | Organizations that focus on type 2 diabetes | Specific focus to make organizations more comparable |
|  | Organizations with a website in English | Understanding the details of the organizations |
|  | Year of founding | Traditional diabetes ecosystem: Organizations that were founded before 31/12/2012.<br>Current diabetes ecosystem: Organizations that were founded after 01/01/2013 |
|  | Traditional diabetes ecosystem: Top 100 based on Revenue | For the traditional diabetes ecosystem based on other articles<sup>40</sup> |
|  | Current diabetes ecosystem: Top 100 based on funding | For the current ecosystem based on related work<sup>39</sup> |
| Exclusion criteria |  | Reason for exclusion |
|  | Companies not generating revenues, or | Identify organizations that are already established in the |
|  | still in the seed stage or series A | market |
|  | Companies with Websites not in English | Understand the analyzed company |

## Appendix C

### – Example of coding

**Table 6.**
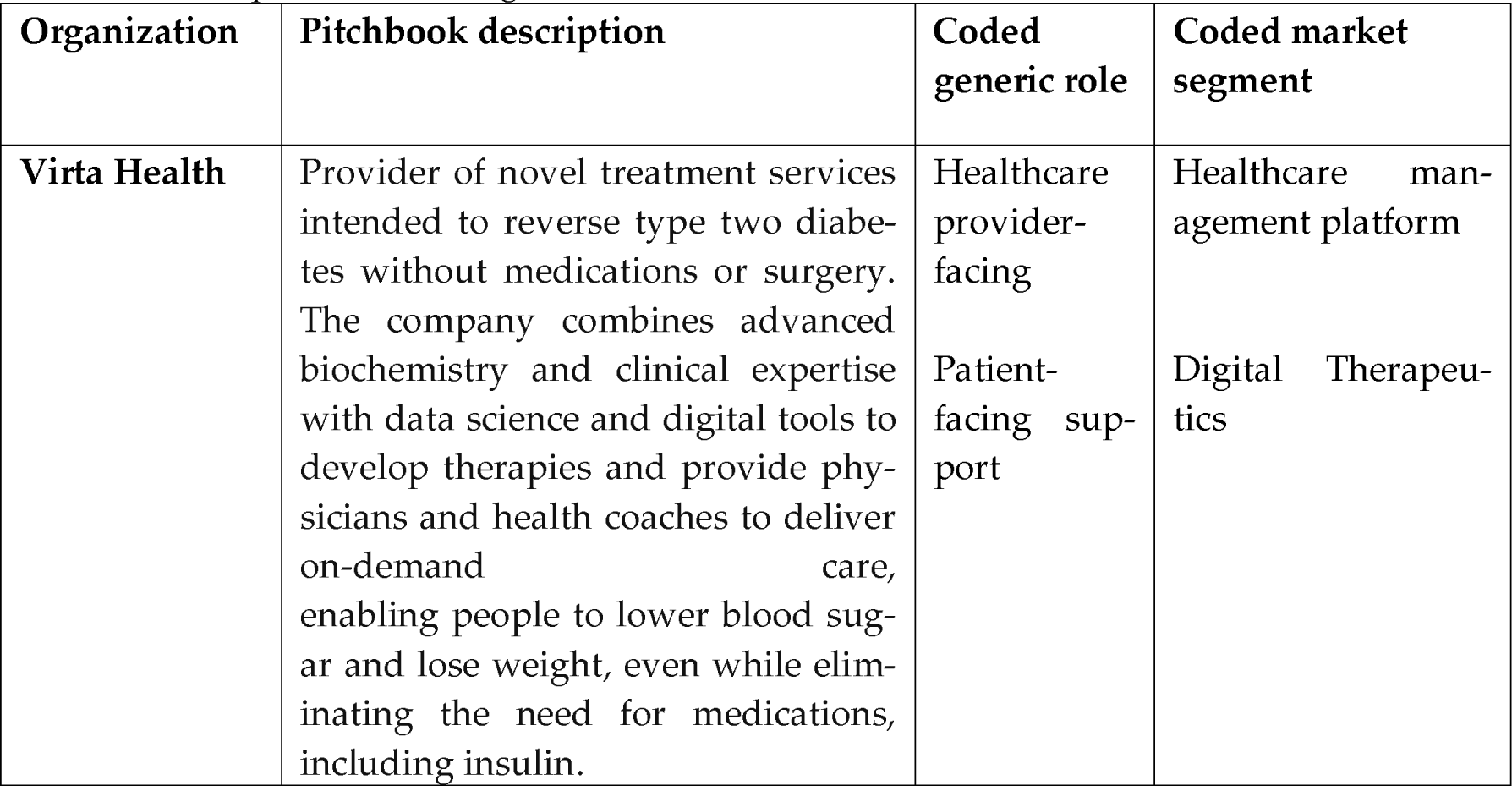
Example of the coding.

| Organization | Pitchbook description | Coded generic role | Coded market segment |
| --- | --- | --- | --- |
| **Virta Health** | Provider of novel treatment services intended to reverse type two diabetes without medications or surgery. The company combines advanced biochemistry and clinical expertise with data science and digital tools to develop therapies and provide physicians and health coaches to deliver on-demand care, enabling people to lower blood sugar and lose weight, even while eliminating the need for medications, including insulin. | Healthcare provider-facing<br>Patient-facing support | Healthcare management platform<br>Digital Therapeutics |

**Table 7.** Example of the description of market segments.

| Market segment and description | Example(s) |
| --- | --- |
| **Digital Therapeutics:**<br>refers to products that deliver medical interventions and therapies. This can be clinical interventions delivered directly to patients via software to treat, manage, or prevent a disease or disorder.<sup>9</sup> | Virta health, Vida Health, Noom |

## Appendix D

### – Expert Interviews

**Table 8.** Overview of the interviews.

| # | Duration (min) | Interviewee's position | Domain |
| --- | --- | --- | --- |
| 1 | 18:13 | Director Alliance Management | Pharmaceutical company |
| 2 | 67:42<br>25:36 | Alliance Manager | Pharmaceutical company |
| 3 | 51:53 | Executive position | Enterprise System and Support |
| 4 | 66:58 | Business development senior manager | Pharmacy |
| 5 | 81:25 | Senior Alliance Manager | Enterprise Systems and |
|  | 106:18 |  | Support |
| 6 | 45:44 | Lead Innovation | Pharmaceutical company |
| 7 | 45:55 | Global Head | Pharmaceutical company |
| 8 | 62:27 | Senior Healthcare Consultant | Consulting company |
| 9 | 66:34 | Head of innovation | Medical Device company |
| 10 | 56:46 | Endocrinologist | Healthcare Provider |

## Appendix E

### – Traditional Ecosystem

**Table 9:**
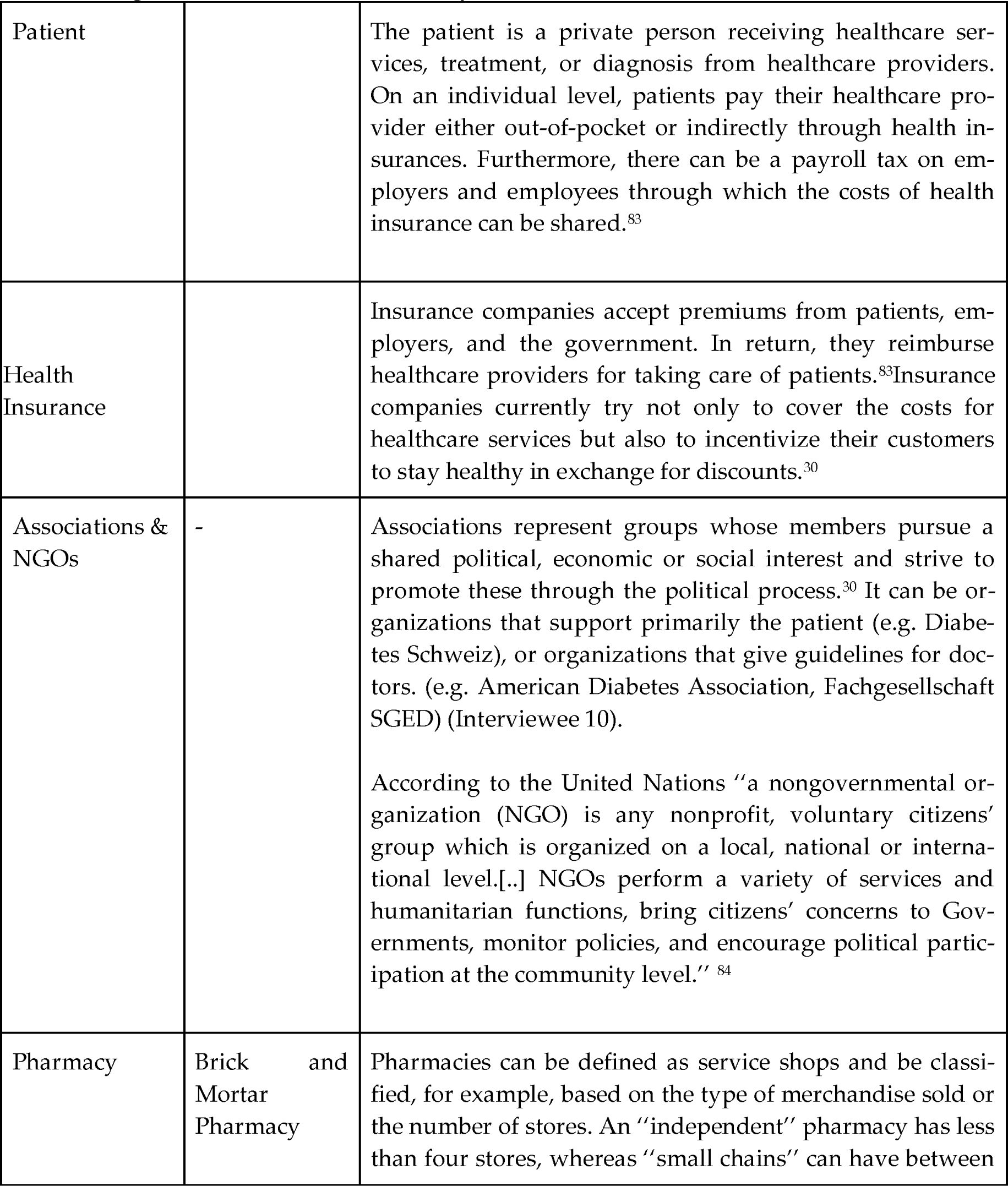
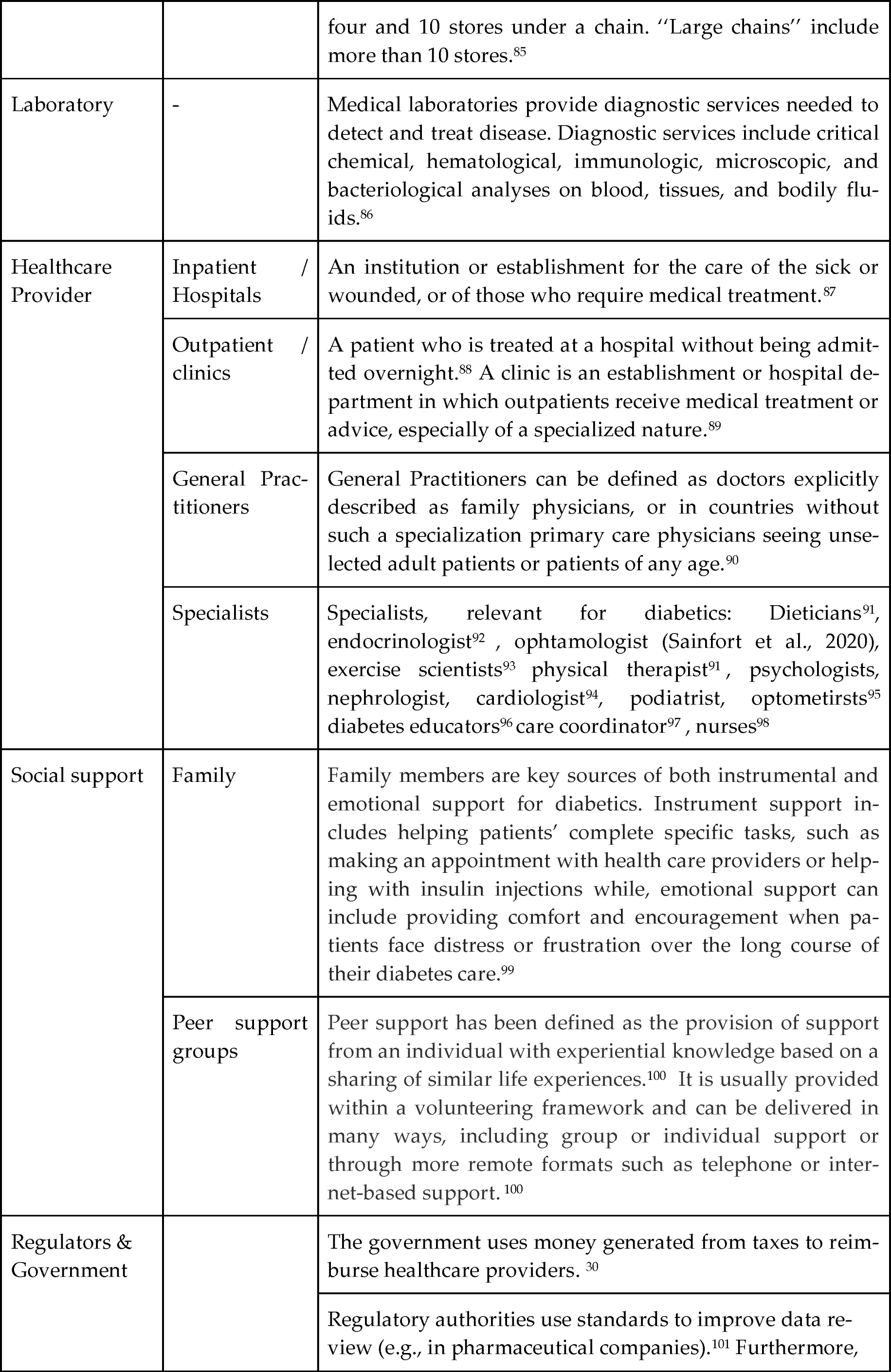
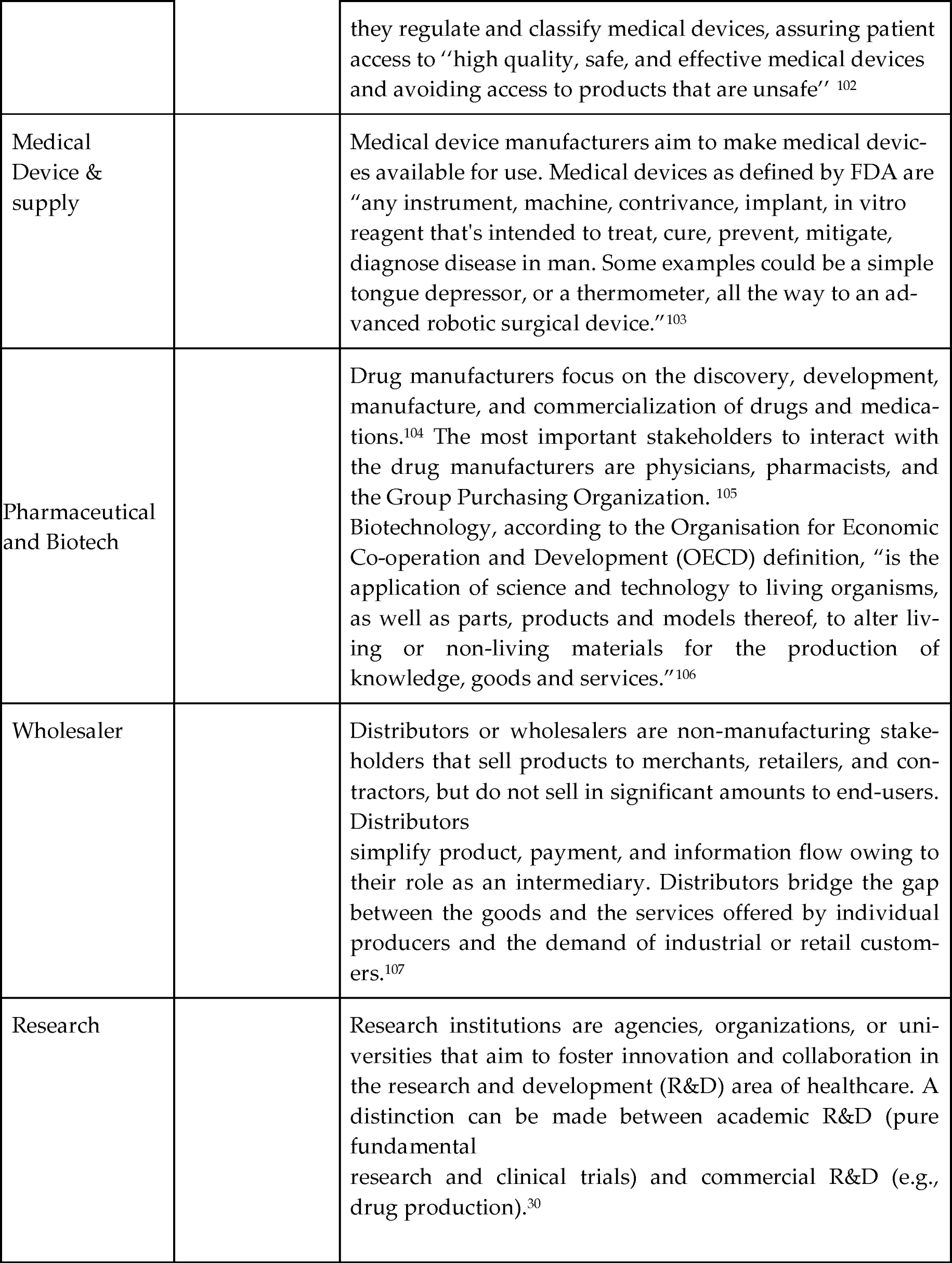
Organizations in the diabetes ecosystem.

## Notes

### Competing Interest Statement

OFG, EP, WM, EF, TK and MJ are affiliated with the Centre for Digital Health Interventions, a joint initiative of the Institute for Implementation Science in Health Care, University of Zurich, the Department of Management, Technology, and Economics at ETH Zurich, and the Institute of Technology Management and School of Medicine at the University of St.Gallen. CDHI is funded in part by CSS, a Swiss health insurer and MavieNext (UNIQA), an Austrian healthcare provider, and MTIP, a Swiss investor company. EF and TK are also a co-founder of Pathmate Technolo-gies, a university spin-off company that creates and delivers digital clinical pathways. However, neither CSS nor Pathmate Technologies, MavieNext or MTIP were involved in the design, analy-sis, or writing of this research.

### Funding Statement

This work was funded in part by the Swiss health insurer CSS.

